# Clarity Plus™ digital PCR: A novel platform for absolute quantification of SARS-CoV-2

**DOI:** 10.1101/2021.05.30.21256718

**Authors:** Shawn Yi Han Tan, Sheng Yi Milton Kwek, Huiyu Low, Yan Ling Joy Pang

## Abstract

In recent years, the usage of digital polymerase chain reaction (dPCR) for various clinical applications has increased exponentially. Considering the growing demand for improved dPCR technology, the Clarity Plus™ dPCR system which features enhanced multiplexing capability and a wider dynamic range for nucleic acid analysis was recently launched. In this study, a dPCR assay optimized for use on Clarity Plus™ was evaluated for the absolute quantification of severe acute respiratory syndrome coronavirus 2 (SARS-CoV-2), the causative agent responsible for the global coronavirus disease 2019 (COVID-19) outbreak. The assay demonstrated good inter- and intra-assay precision, accuracy, as well as excellent linearity across a range of over 6 orders of magnitude for target gene quantification. In addition, comparison of the assay on both dPCR and qPCR platforms revealed that dPCR exhibited a slightly higher sensitivity compared to its qPCR counterpart when quantifying SARS-CoV-2 at a lower concentration. Overall, the results showed that the dPCR assay is a reliable and effective approach for the absolute quantification of SARS-CoV-2 and can potentially be adopted as a molecular tool for detecting low viral load in patients and wastewater surveillance of COVID-19.

## 1. Introduction

Digital Polymerase Chain Reaction (dPCR) was first introduced by Volgelstein and Kinzler in a 1999 publication in which they utilized a series of four 96 well plates to physically partition samples obtained from colorectal cancer patients in order to detect mutations in the *ras* oncogene [1]. Being the third generation PCR platform, dPCR works by separating a PCR mixture into many independent nanoliter sub-reactions, leading to individual partitions having either a few or no target sequences. Upon completion of PCR, the proportion of positive and negative partitions for each sample is determined and used to compute the absolute nucleic acid copy number using Poisson statistics [2]. The nanoliter sub-reaction volume in each partition causes increased congregation of the PCR mix, reducing template competition as well as sample susceptibility to PCR inhibitors, this gives dPCR assays increased relative sensitivity to detect rare mutations in a large background of various wild-type sequences [3]. For example, in the case of detecting rare circulating tumor DNA, dPCR provides a higher sensitivity of up to 0.001% without compromising accuracy [2, 4]. In comparison to traditional PCR and qPCR, the lack of dependency on standard curves for reference enables dPCR to perform absolute end-point detection and quantification of desired nucleic acid targets with increased precision and accuracy [3, 5].

Commercial dPCR platforms that are available in the market mainly employ two different methods of partitioning. The first method involves the generation of water-oil emulsion droplets. This technology is employed by the QX100™/200™ droplet digital PCR system (Bio-Rad Laboratories) and the Naica® System (Stilla Technologies). The second method is a chip-based dPCR technology used by BioMark™ HD (Fluidigm), QuantStudio® 3D (Thermo Fischer Scientific), and Clarity™ (JN Medsys) [2, 6]. Since the mid-2000s, a plethora of new dPCR applications have been developed which consequently led to an exponential increase in the number of scientific paper published [7]. A quick search on NCBI with “digital PCR” as the keyword returned a search result of close to 4,000 publications. With the increased demand for dPCR in research and clinical diagnostics, the Clarity Plus™ system was recently launched which features improved multiplexing capability and a wider dynamic range for analysis as compared to its predecessor, the Clarity™ system. The platform features a chip-in-a-tube technology which combines the advantages from both droplet and chip-based dPCR, bringing about higher sensitivity, robustness, reliability, as well as accuracy for nucleic acid detection and quantification. The use of Clarity™ system had been reported in various applications such as detection of isoniazid heteroresistance in *Mycobacterium tuberculosis*, the monitoring of hematologic malignancies in clinical studies, and Epstein-Barr virus (EBV) viral load quantification for nasopharyngeal carcinoma [8-10].

The severe acute respiratory syndrome coronavirus 2 (SARS-CoV-2) was first reported in Wuhan, China, at the end of 2019. This virus is highly transmissible and has caused a global pandemic of acute respiratory disease which was subsequently named Coronavirus Disease 2019 (COVID-19). As of May 2021, over 160 million COVID-19 cases and 3.5 million deaths have been reported worldwide [11]. This pandemic has posed a severe threat to global public health which necessitates the development of reliable tests for sensitive and accurate detection of SARS-CoV-2. The SARS-CoV-2 genome is made up of a single linear positive strand RNA segment of approximately 30,000 bases comprising of ORF1a, ORF1b, S, E, M, RNA-dependent RNA Polymerase (RdRP), N1, and N2 sequences [12, 13]. Presently, reverse transcription-quantitative polymerase chain reaction (RT-qPCR) is the gold standard method for the diagnosis of COVID-19 worldwide [14]. Various RT-qPCR assays have been developed worldwide which detect SARS-CoV-2 RNA through amplification of 2 or 3 distinct segments [15]. However, there have been reports of false negative cases which have adverse implications for prompt isolation of positive cases and management of the disease. False negative results can be attributed to different factors including low viral load, testing too early in the course of infections and low analytical sensitivity [5, 16, 17]. In light of these challenges, other methodologies including dPCR have been explored for SARS-CoV-2 detection to complement the currently available tests [18]. In this study, a TaqMan-based dPCR assay optimized on the Clarity Plus™ system was evaluated for SARS-CoV-2 detection and the results demonstrate that this technology can potentially be employed in clinical settings for detection and absolute quantification of low copy viral load.

## 2. Materials & Methods

### 2.1 Reagent preparations

Each 15μl Clarity Plus™ COVID-19 reverse transcription (RT) - dPCR reaction mix consisted of 7.5μl Clarity Plus™ COVID-19 Probe RT-dPCR Mastermix (2x), 0.75μl Clarity Plus™ COVID-19 RT Mix (20x), 0.75μl Clarity Plus™ JN solution (20x), 1μl Clarity Plus™ COVID-19 Primer & Probe Mix (N1-FAM, ORF1ab-Quasar670 and human RNaseP-HEX) and 5μl RNA Sample/Positive control/ NTC following manufacturer’s instructions (JN Medsys, Cat no. 10028). Clarity Plus™ JN solution is a proprietary formula optimized for robust performance on Clarity Plus™ high density chips. The prepared mix was placed in the thermocycler for reverse transcription at 55°C for 15mins and a continuous 4°C step till the sample is ready for loading.

Using the Clarity™ auto loader, the mix was delivered onto the chip where it was sub-divided into 40,000 partitions and subsequently sealed with Clarity™ Seal Enhancer with addition of 245μl Clarity™ Sealing fluid (Fig 1). All sealed chips underwent another round of thermal cycling using the following parameters: Initial 1 cycle of 95 °C for 5 min and 40 cycles of 95°C for 50s, 55°C for 90s, and 70°C for 5 min (ramp rate = 2.5 °C/s). After PCR amplification, the tube strips were transferred to the Clarity Plus™ Reader which was set to detect the N1 gene on FAM channel for the detection of FAM fluorescence and ORF1ab gene on Cy5 channel for the detection of Quasar670 fluorescence. The data was analyzed with the Clarity™ software (version 4.1), and a proprietary algorithm was used for setting individual thresholds based on fluorescent intensities to determine the proportion of positive partitions. Using this information, the software determines the DNA copies per microliter of dPCR mix using Poisson statistics. The mean partition volume of 0.31 nl was used for copy number calculation.

**Fig 1:**
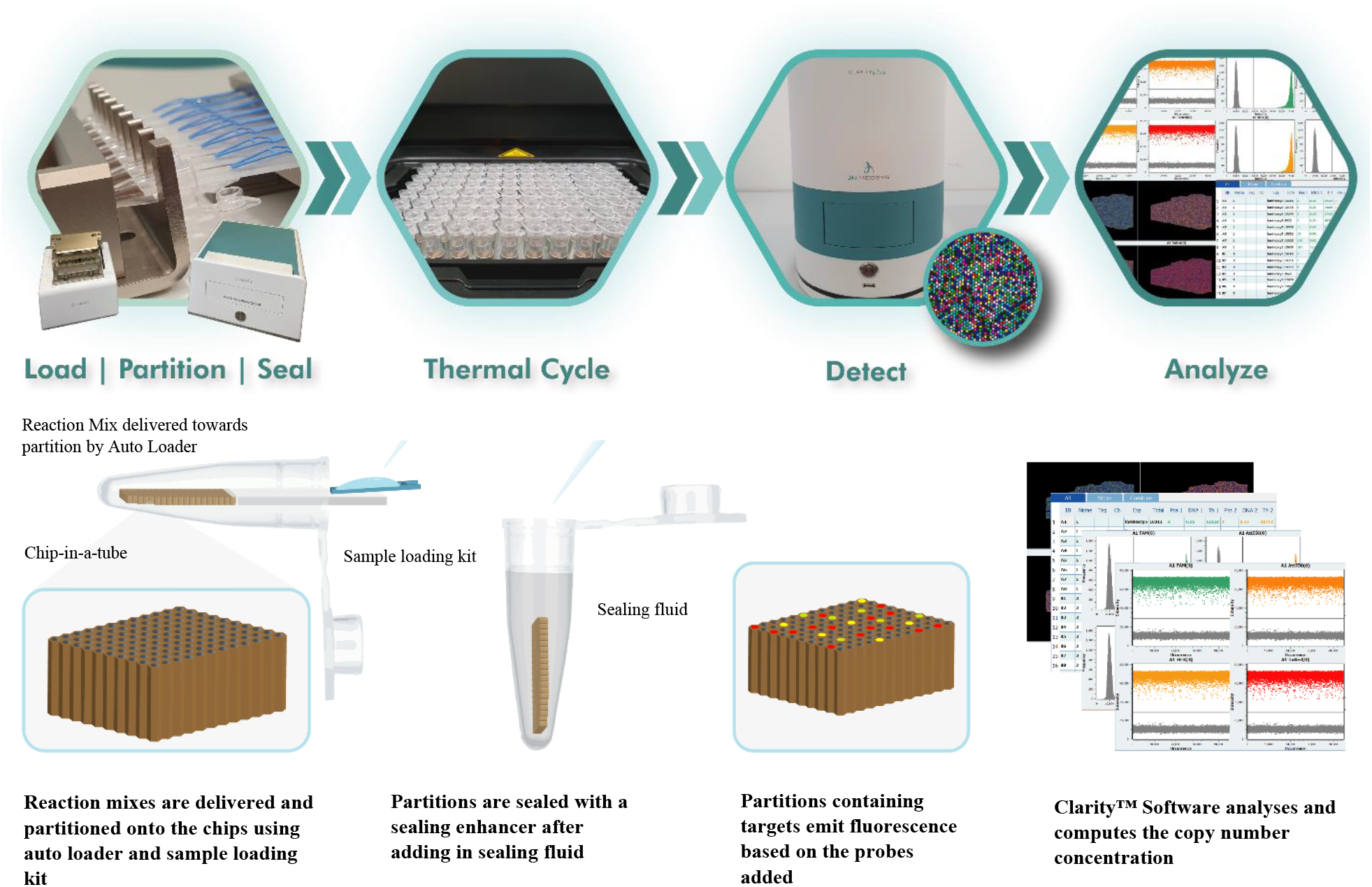
Experimental workflow of Clarity Plus™ digital PCR. Similar Clarity™, the Clarity Plus™ system adopts a unique chip-in-a-tube format which allows digital PCR to be performed with ease and speed via simultaneous loading and partitioning of eight reaction mixes using an auto loader. The reaction mixes are subsequently sealed in the partitions with the sealing enhancer and filled with sealing fluid. Following which, the reactions are subjected to thermal cycling for PCR amplification. After amplification, the tube-strips are transferred to the Clarity Plus™ reader where fluorescence signals in respective partitions are detected simultaneously. Lastly, the proportion of positive to negative partitions is analyzed by the Clarity™ software (version 4.1) to produce the copy number concentration of each sample using Poisson statistics.

### 2.2 Quantification of N1 Synthetic RNA

N1 and ORF1ab synthetic RNA (GenScript) was quantified using the first generation Clarity™ system (JN Medsys) [6]. The stock was serially diluted based on theoretical copies per microliter formula^a^. Upon identifying the quantifiable range within the dilutions, the counts were readjusted and reconfirmed with Clarity™. Copy number validation on Clarity Plus™ was subsequently obtained by running readjusted copy number on both Clarity™ and Clarity Plus™ systems simultaneously (data not shown).

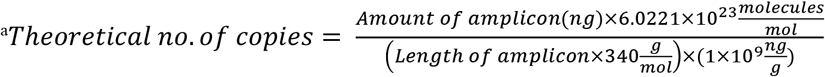

### 2.3 SARS-CoV-2 linearity study and limit of quantification (LOQ)

The Linearity and LOQ of SARS-CoV-2 RT-dPCR assay were obtained by quantifying a single copy of N1 and ORF1ab gene obtained from a series of serial dilutions with a range of expected target concentration of 0.02 to 30,000 copies/μl reaction mixture. The dilutions were yielded from a theoretical 3×10^12^ copies/µl stock. N1 and ORF1ab probes and primers used were custom designed in JN Medsys. All copy number were previously validated on Clarity™ systems before dilution. A total of three independent experimental runs were performed with the same concentration range in duplicates.

The quantification limit for SARS-CoV-2 RT-dPCR assay on Clarity Plus™ was obtained by serially diluting a single copy N1 and ORF1ab gene with expected concentrations ranging from 0.125 to 1 copies/μl reaction mixture. The dilutions were yielded from the same stock and three independent LOQ runs were performed with respective concentrations in duplicates.

### 2.4 SARS-CoV-2 Assay limit of detection (LOD) on inactivated SARS-CoV-2 virus

AccuPlex™ SARS-CoV-2 RNA (Seracare #0505-0126) was first extracted using EX3600 Automated Nucleic Acid Extraction System (Liferiver™) according to manufacturer’s instructions. The extracted RNA stock was subsequently quantified with Clarity Plus™. To obtain preliminary LOD for extracted RNA in SARS-CoV-2 dPCR assay, the RNA stock was serially diluted 2, 4, 8, 16, and 32 times and quantified in triplicates respectively.

The sensitivity of the N1 SARS-CoV-2 RT-dPCR assay was evaluated on both Clarity Plus™ (JN Medsys) dPCR and the QuantStudio® 3 qPCR (Thermo Fisher Scientific) platforms. As QuantStudio® 3 qPCR system is unable to read the Quasar670 fluorescence, only ORF1ab digital PCR data is provided. The extracted RNA stock was diluted 8 and 16 times to achieve an expected concentration of 0.225 and 0.1125 copies/μl respectively. Reagent preparations and Clarity Plus™ workflow for SARS-CoV-2 RT-dPCR assay remained unchanged. For comparison with SARS-CoV-2 RT-qPCR assay, each 20μl reaction mix consisted of 10μl Clarity Plus™ COVID-19 Probe RT-dPCR Mastermix (2x), 1μl Clarity Plus™ COVID-19 RT Mix (20x), 1.33μl Clarity Plus™ COVID-19 Primer & Probe Mix, 5μl RNA Sample/Positive control/ NTC) and topped up with nuclease free water. The samples were loaded onto a 96 well plate before being amplified on QuantStudio® 3 real-time qPCR machine. All samples underwent thermal cycling using the following parameters: Initial cycle of 55°C for 15 min, 95°C for 2 min, 40 cycles of 95°C for 3s, and 55°C for 30s (ramp rate 1.6 °C/s). Upon completion, the data captured by QuantStudio® 3 were further analyzed with the Design and Analysis software (version 2.4.3) by Thermo Fisher Scientific.

### 2.6 Statistical Analysis

All linear regression analysis on data obtained from Clarity Plus™ was performed on GraphPad Prism® (version 8.0.1).

## 3. Results and Discussion

### 3.1 Absolute quantification of SARS-CoV-2 target using Clarity Plus™ system

The linearity and the limit of quantification (LOQ) for the Clarity Plus™ SARS-CoV-2 RT-dPCR assay were first determined using N1 and ORF1ab Synthetic RNA (GenScript). A series of 10 serially diluted RNA aliquots ranging from 0.02 to 30,000 copies/μl was obtained from a 3×10^12^ copies/μl stock. Three independent experiments were conducted to quantify these RNA aliquots using the Clarity Plus™ system. The average measured concentration from three independent runs is shown in Table 1a for N1 and Table 1b for ORF1ab. The measured concentrations of N1 and ORF1ab were found to be closely associated with their expected concentrations except for two concentrations at 0.02 and 30,000 copies/μl respectively. When measured concentrations (0.2 to 20,000 copies/μl) was plotted against their expected values, linear regression analysis revealed a R^2^ value of 0.998 for both N1 (Fig 2a) and ORF1ab (Fig 2b) across a dynamic range of over 6 orders of magnitude. This indicates that excellent linearity is possible across a dynamic range of 0.2 to 20,000 copies/μl.

**Table 1a.**
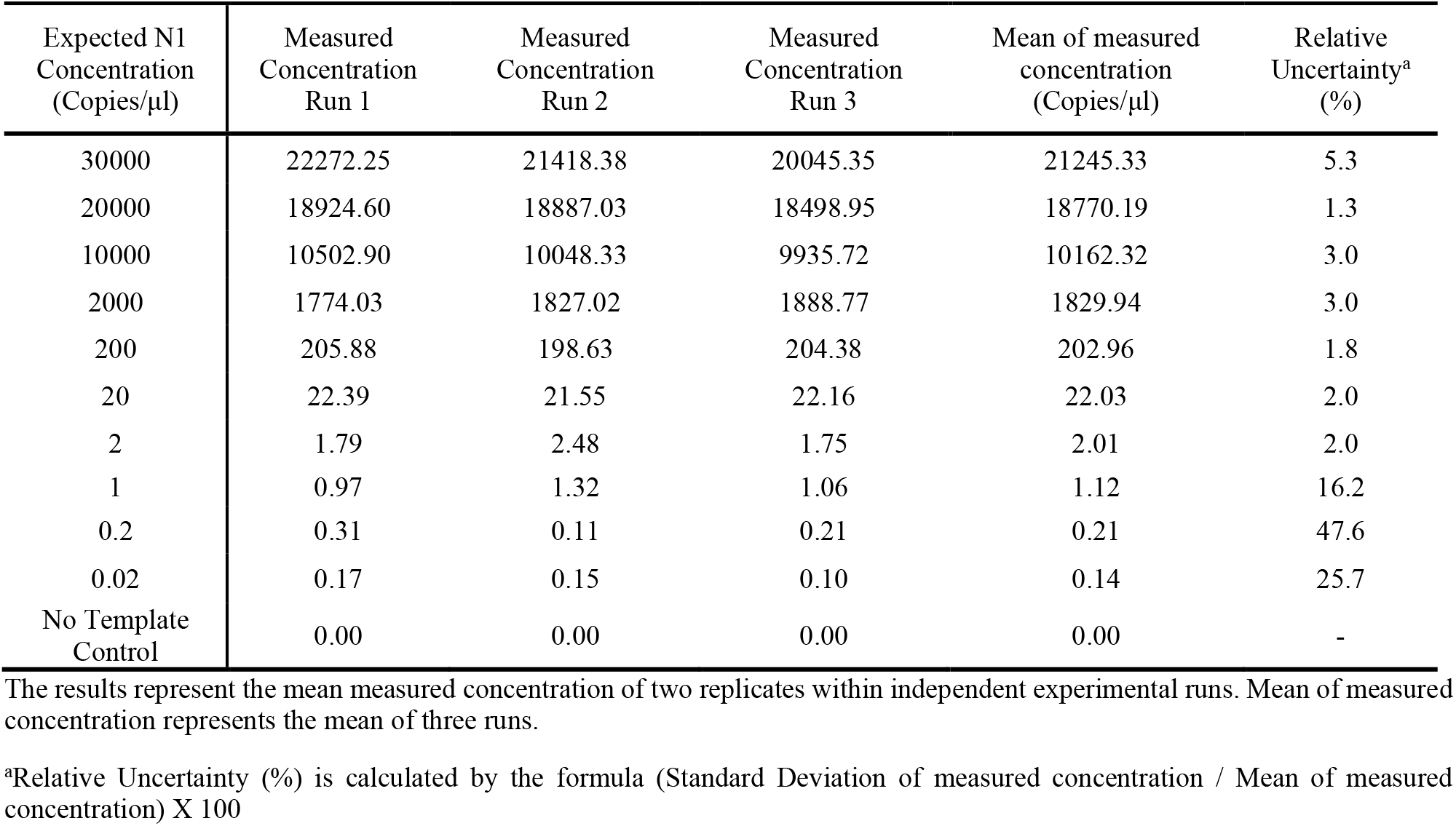
SARS-CoV-2 RT-dPCR assay linearity study on Clarity Plus™ with three independent experimental runs, using a series of expected N1 concentrations ranging from 0.02 to 30,000 copies/μl.

| Expected N1 Concentration (Copies/μl) | Measured Concentration Run 1 | Measured Concentration Run 2 | Measured Concentration Run 3 | Mean of measured concentration (Copies/μl) | Relative Uncertainty<sup>a</sup> (%) |
| --- | --- | --- | --- | --- | --- |
| 30000 | 22272.25 | 21418.38 | 20045.35 | 21245.33 | 5.3 |
| 20000 | 18924.60 | 18887.03 | 18498.95 | 18770.19 | 1.3 |
| 10000 | 10502.90 | 10048.33 | 9935.72 | 10162.32 | 3.0 |
| 2000 | 1774.03 | 1827.02 | 1888.77 | 1829.94 | 3.0 |
| 200 | 205.88 | 198.63 | 204.38 | 202.96 | 1.8 |
| 20 | 22.39 | 21.55 | 22.16 | 22.03 | 2.0 |
| 2 | 1.79 | 2.48 | 1.75 | 2.01 | 2.0 |
| 1 | 0.97 | 1.32 | 1.06 | 1.12 | 16.2 |
| 0.2 | 0.31 | 0.11 | 0.21 | 0.21 | 47.6 |
| 0.02 | 0.17 | 0.15 | 0.10 | 0.14 | 25.7 |
| No Template Control | 0.00 | 0.00 | 0.00 | 0.00 | - |

The results represent the mean measured concentration of two replicates within independent experimental runs. Mean of measured concentration represents the mean of three runs.
<sup>a</sup>Relative Uncertainty (%) is calculated by the formula (Standard Deviation of measured concentration / Mean of measured concentration) X 100

**Table 1b.**
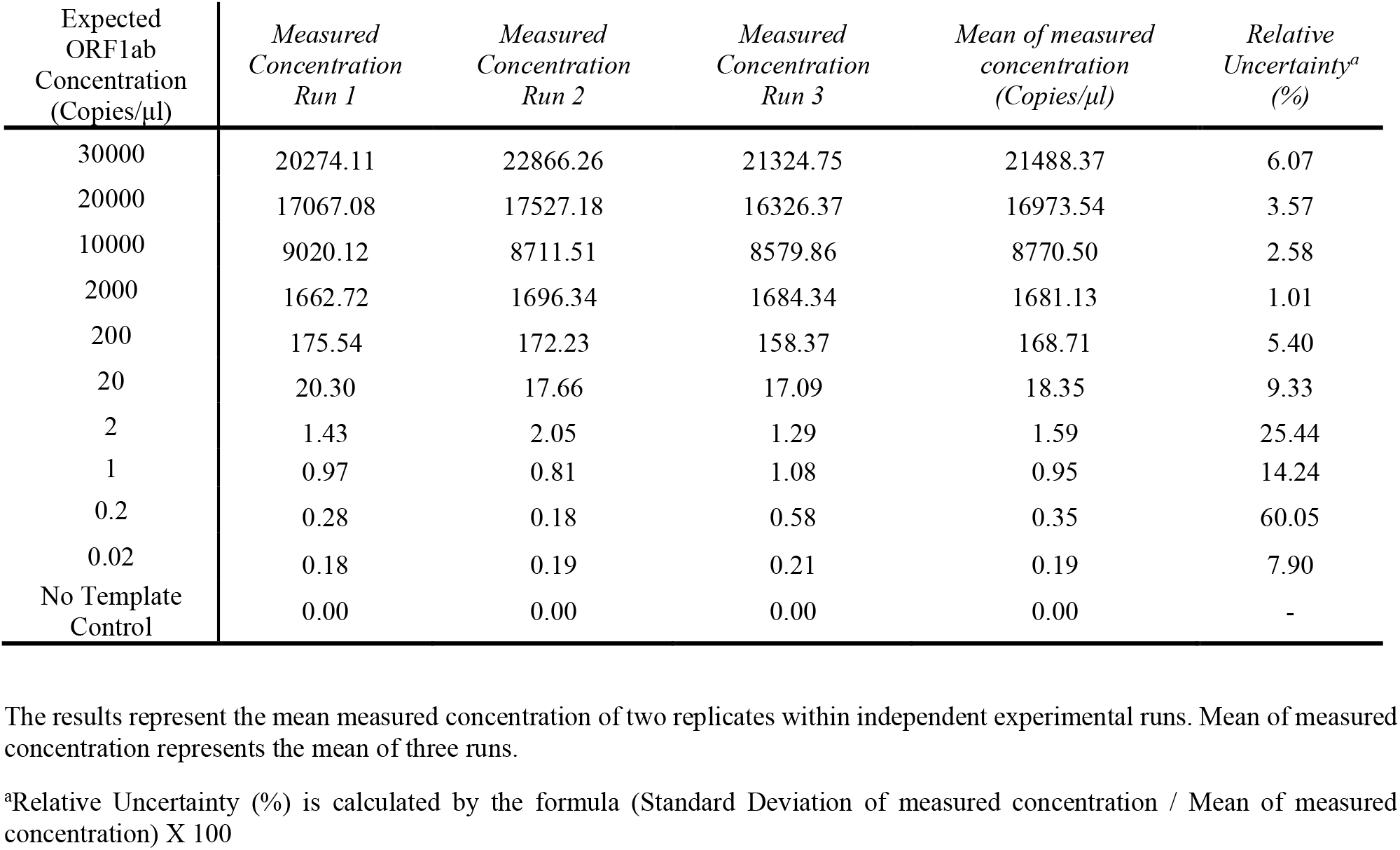
SARS-CoV-2 RT-dPCR assay linearity study on Clarity Plus™ with three independent experimental runs, using a series of expected ORF1ab concentrations ranging from 0.02 to 30,000 copies/μl.

| Expected ORF1ab Concentration (Copies/μl) | Measured Concentration Run 1 | Measured Concentration Run 2 | Measured Concentration Run 3 | Mean of measured concentration (Copies/μl) | Relative Uncertainty<sup>a</sup> (%) |
| --- | --- | --- | --- | --- | --- |
| 30000 | 20274.11 | 22866.26 | 21324.75 | 21488.37 | 6.07 |
| 20000 | 17067.08 | 17527.18 | 16326.37 | 16973.54 | 3.57 |
| 10000 | 9020.12 | 8711.51 | 8579.86 | 8770.50 | 2.58 |
| 2000 | 1662.72 | 1696.34 | 1684.34 | 1681.13 | 1.01 |
| 200 | 175.54 | 172.23 | 158.37 | 168.71 | 5.40 |
| 20 | 20.30 | 17.66 | 17.09 | 18.35 | 9.33 |
| 2 | 1.43 | 2.05 | 1.29 | 1.59 | 25.44 |
| 1 | 0.97 | 0.81 | 1.08 | 0.95 | 14.24 |
| 0.2 | 0.28 | 0.18 | 0.58 | 0.35 | 60.05 |
| 0.02 | 0.18 | 0.19 | 0.21 | 0.19 | 7.90 |
| No Template Control | 0.00 | 0.00 | 0.00 | 0.00 | - |

The results represent the mean measured concentration of two replicates within independent experimental runs. Mean of measured concentration represents the mean of three runs.
<sup>a</sup>Relative Uncertainty (%) is calculated by the formula (Standard Deviation of measured concentration / Mean of measured concentration) X 100

**Fig 2:**
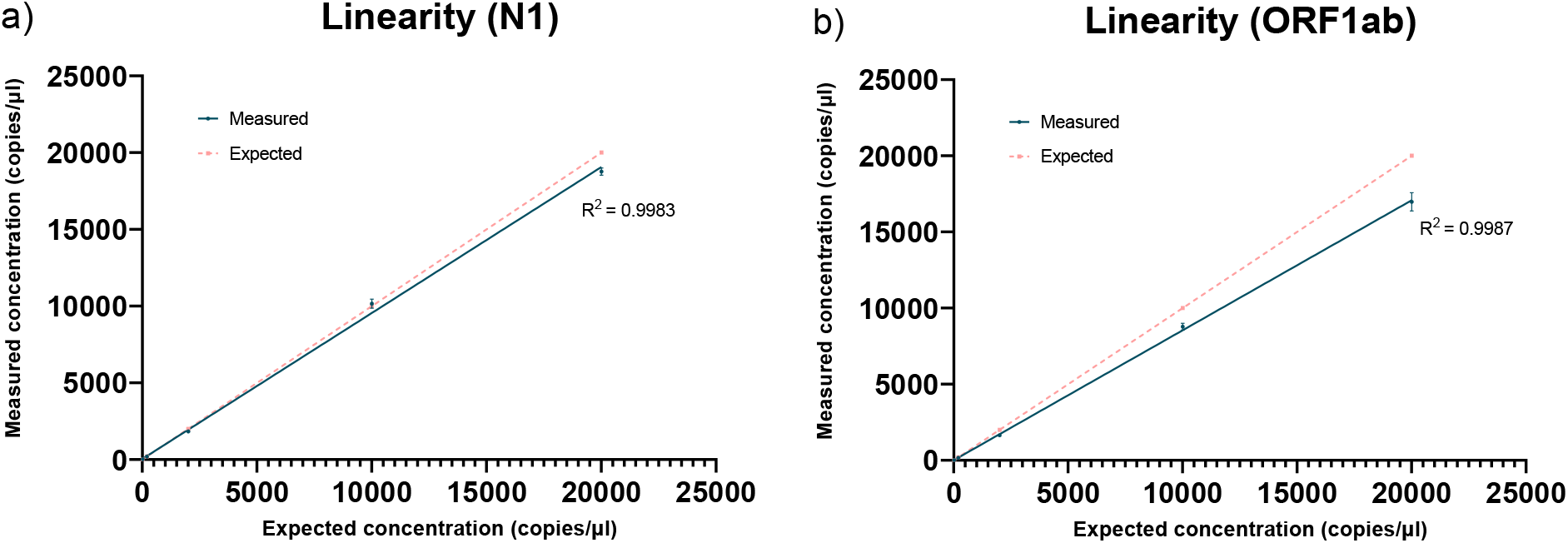
SARS-CoV-2 RT-dPCR Assay linearity study with Clarity Plus™ using a series of serially diluted N1 synthetic RNA. a) Figure of N1 linearity b) Figure of ORF1ab linearity

Inter and intra-assay precision for the SARS-CoV-2 RT-dPCR assay using Clarity Plus™ were also evaluated based on relative uncertainty (RU) between data values. As summarized in Table 1, the mean measured N1 and ORF1ab concentrations between three independent runs are well correlated with a RU of less than 10% except for the 4 lowest dilutions (0.02, 0.2, 1, and 2 copies/µl). Within the respective assays (Table 2a and 2b), the data of N1 and ORF1ab shows a good intra-assay precision between duplicate values of respective concentrations (<10% RU) except those from lower dilutions (0.02, 0.2, 1, 2 copies/μl). High RU values at lower concentrations does not accurately portray the relationship between data since a minor variation between independent runs such as 0.1 and 0.2 copies/μl will cause a significant 50% increase in RU. Furthermore, linear regression analysis on lower concentrations of 0.2, 1, and 2 copies/μl had a R^2^ value of 0.915 for N1 and 0.835 for ORF1ab which indicates a good linearity as well as close association between mean measured concentration and expected concentration despite having high RU values (Fig 3).

**Table 2a.** SARS-CoV-2 RT-dPCR assay linearity study on Clarity Plus™ with three independent experimental runs in duplicates, using a series of expected N1 concentrations ranging from 0.02 to 30,000 copies/μl

| Expected Concentration (Copies/μl) | Measured Concentration Run 1 |  | Relative Uncertainty<sup>a</sup> (%) | Measured Concentration Run 2 |  | Relative Uncertainty<sup>a</sup> (%) | Measured Concentration Run 3 |  | Relative Uncertainty<sup>a</sup> (%) |
| --- | --- | --- | --- | --- | --- | --- | --- | --- | --- |
| 30000 | 23590.46 | 20954.04 | 8.37 | 22618.00 | 20218.76 | 7.92 | 19069.93 | 21020.77 | 6.88 |
| 20000 | 19149.16 | 18700.05 | 1.68 | 18074.79 | 19699.26 | 6.08 | 18079.42 | 18918.48 | 3.21 |
| 10000 | 10409.95 | 10595.85 | 1.25 | 10081.11 | 10015.54 | 0.46 | 10295.95 | 9575.48 | 5.13 |
| 2000 | 1792.33 | 1755.73 | 1.46 | 1809.06 | 1844.98 | 1.39 | 1817.38 | 1960.16 | 5.35 |
| 200 | 207.07 | 204.70 | 0.81 | 198.54 | 198.72 | 0.06 | 212.81 | 195.96 | 5.83 |
| 20 | 22.56 | 22.23 | 1.04 | 20.80 | 22.30 | 4.92 | 21.17 | 22.90 | 5.55 |
| 2 | 1.91 | 1.68 | 9.06 | 2.82 | 2.15 | 19.06 | 1.42 | 2.08 | 26.67 |
| 1 | 0.97 | 0.97 | 0 | 1.51 | 1.13 | 20.36 | 1.06 | 1.06 | 0 |
| 0.2 | 0.41 | 0.21 | 45.62 | 0.15 | 0.07 | 51.43 | 0.20 | 0.21 | 3.45 |
| 0.02 | 0.14 | 0.21 | 28.28 | 0.07 | 0.23 | 75.42 | 0.13 | 0.07 | 42.43 |
| No Template Control | 0.00 | 0.00 | - | 0.00 | 0.00 | - | 0.00 | 0.00 | - |

The results represent the individual measured concentration of two replicates within independent experimental runs.
<sup>a</sup>Relative Uncertainty (%) is calculated by the formula (Standard Deviation of measured concentration / Mean of measured concentration) X 100

**Table 2b.** SARS-CoV-2 RT-dPCR assay linearity study on Clarity Plus™ with three independent experimental runs in duplicates, using a series of expected ORF1ab concentrations ranging from 0.02 to 30,000 copies/μl

| Expected Concentration (Copies/μl) | Measured Concentration Run 1 |  | Relative Uncertainty<sup>a</sup> (%) | Measured Concentration Run 2 |  | Relative Uncertainty<sup>a</sup> (%) | Measured Concentration Run 3 |  | Relative Uncertainty<sup>a</sup> (%) |
| --- | --- | --- | --- | --- | --- | --- | --- | --- | --- |
| 30000 | 23295.77 | 17252.46 | 21.08 | 23599.31 | 22133.22 | 4.53 | 21493.69 | 21155.82 | 1.12 |
| 20000 | 16655.78 | 17478.38 | 3.41 | 17613.36 | 17441.00 | 0.70 | 16061.96 | 16590.78 | 2.29 |
| 10000 | 9005.66 | 9034.58 | 0.23 | 8690.26 | 8732.76 | 0.34 | 8452.98 | 8706.74 | 2.09 |
| 2000 | 1644.24 | 1681.20 | 1.57 | 1780.67 | 1612.01 | 7.03 | 1684.34 | 1684.33 | 0.00 |
| 200 | 176.66 | 174.42 | 0.90 | 176.43 | 168.02 | 3.45 | 153.89 | 162.85 | 4.00 |
| 20 | 19.67 | 20.93 | 4.39 | 18.70 | 16.63 | 8.29 | 18.12 | 16.06 | 8.52 |
| 2 | 1.30 | 1.55 | 12.41 | 2.08 | 2.02 | 2.07 | 1.11 | 1.48 | 20.20 |
| 1 | 1.19 | 0.76 | 31.19 | 0.42 | 1.20 | 68.09 | 0.98 | 1.19 | 13.69 |
| 0.2 | 0.21 | 0.34 | 33.43 | 0.07 | 0.30 | 87.91 | 0.14 | 1.02 | 107.29 |
| 0.02 | 0.07 | 0.28 | 84.85 | 0.16 | 0.22 | 22.33 | 0.42 | 0.00 | 141.42 |
| No Template Control | 0.00 | 0.00 | - | 0.00 | 0.00 | - | 0.00 | 0.00 | - |

The results represent the individual measured concentration of two replicates within independent experimental runs.
<sup>a</sup>Relative Uncertainty (%) is calculated by the formula (Standard Deviation of measured concentration / Mean of measured concentration) X 100

**Fig 3:**
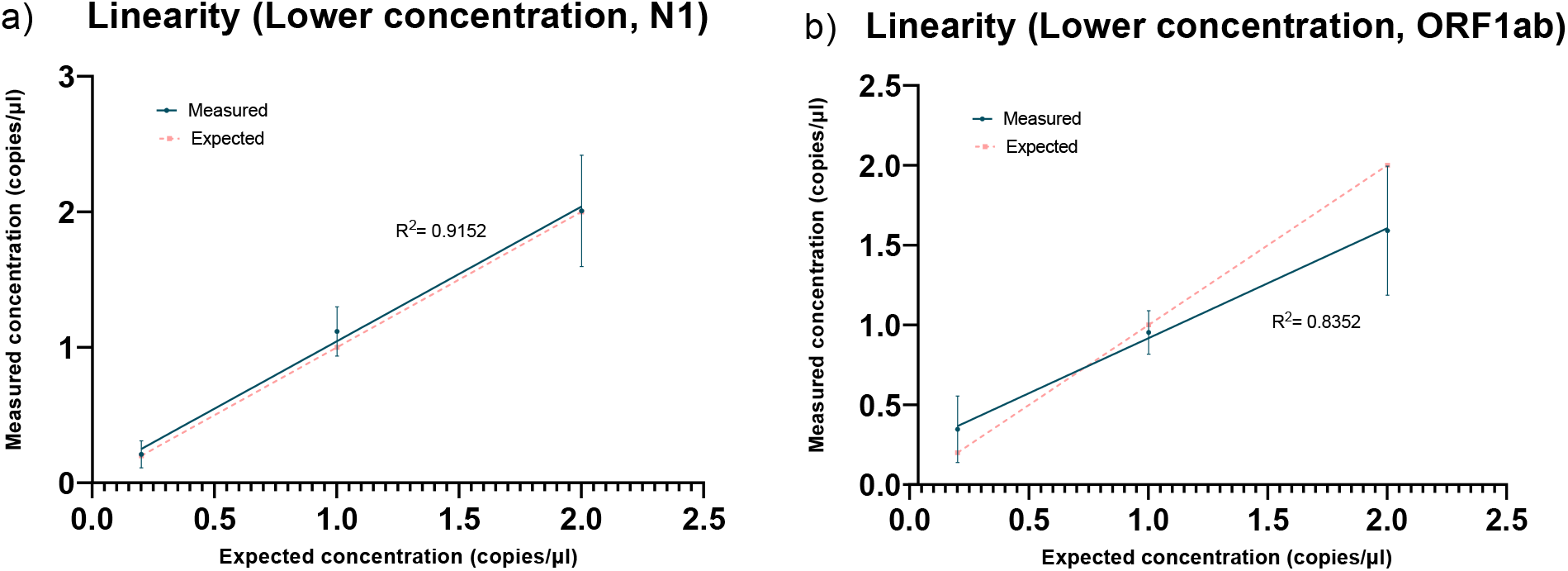
SARS-CoV-2 RT-dPCR Assay linearity study at lower concentrations of 0.2, 1, and 2 copies/μl. a) Figure of N1 linearity at lower concentrations b) Figure of ORF1ab linearity at lower concentrations

### 3.2 Limit of quantification for SARS-CoV-2 RT-dPCR assay

The quantification limit of SARS-CoV-2 RT-dPCR assay was established with a series of four expected N1 and ORF1ab synthetic RNA concentrations of 0.125 to 1 copies/μl serially diluted from a stock RNA solution. Three independent experiments were conducted to establish limit of quantification for the assay with Clarity Plus™. Despite having high RU values, mean measured concentrations of 1, 0.5, and 0.25 copies/μl portrayed a close association with their expected concentration as linear regression analysis on these concentrations revealed a strong linear correlation with a R^2^ value of 0.98 for N1 (Fig 4a) and 0.83 for ORF1ab (Fig 4b). The quantification limit of the assay was determined to be 0.25 copies/μl as mean measured concentration at 0.125 copies/μl does not mimic the linear relationship of a 2-fold dilution (Table 3a and b).

**Fig 4:**
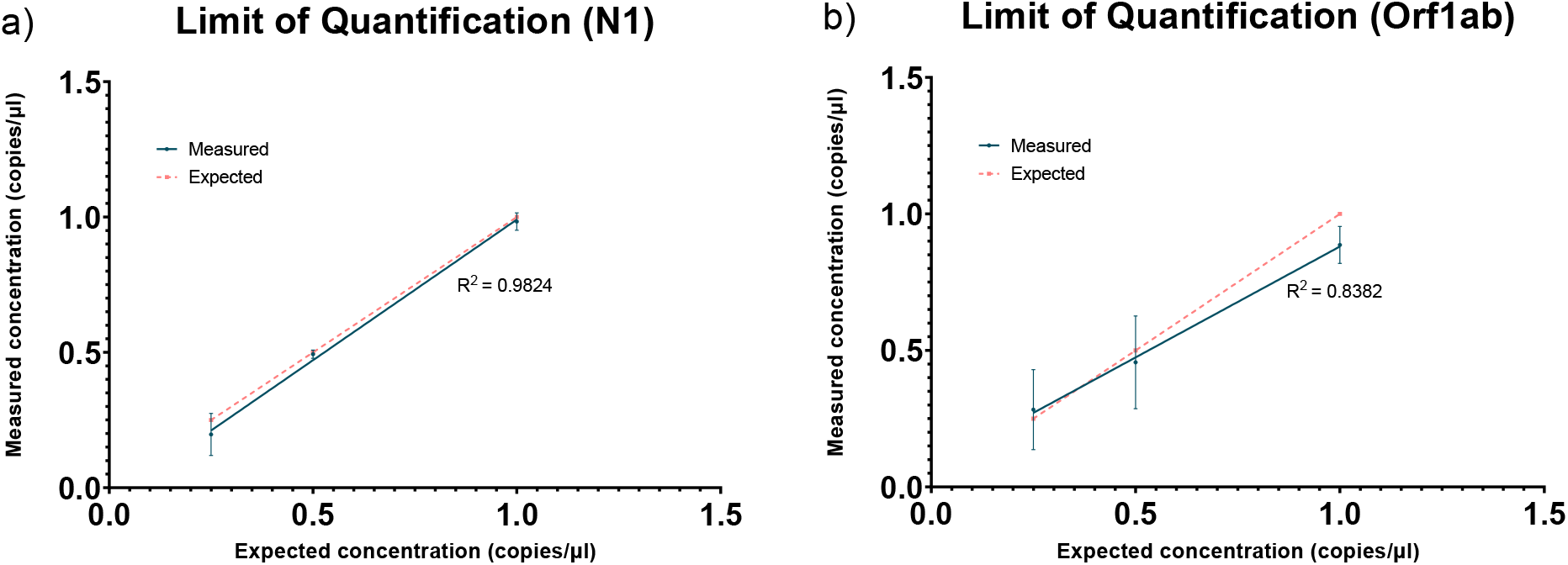
SARS-CoV-2 RT-dPCR Assay limit of quantification by Clarity Plus™ using a series of serially diluted N1 Synthetic RNA. a) Figure of N1 limit of quantification b) Figure of ORF1ab limit of quantification

**Table 3a.** Limit of Quantification (LOQ) of N1 SARS-CoV-2 RT-dPCR assay quantified using Clarity Plus™. Three independent experimental runs were performed with a series of expected N1 concentrations ranging from 0.125 to 1 copies/μl.

| Expected Concentration (Copies/μl) | Measured Concentration Run 1 | Measured Concentration Run 2 | Measured Concentration Run 3 | Mean of measured concentration (Copies/μl) | Relative Uncertainty<sup>a</sup> (%) |
| --- | --- | --- | --- | --- | --- |
| 1 | 0.97 | 1.02 | 0.96 | 0.98 | 3.2 |
| 0.5 | 0.49 | 0.51 | 0.48 | 0.49 | 3.1 |
| 0.25 | 0.26 | 0.11 | 0.22 | 0.20 | 39.4 |
| 0.125 | 0.23 | 0.19 | 0.18 | 0.20 | 13.2 |
| No Template Control | 0.00 | 0.00 | 0.00 | 0.00 | - |

The results represent the mean measured concentration of two replicates within independent experimental runs. Mean of measured concentration represents the mean of three runs.
<sup>a</sup>Relative Uncertainty (%) is calculated by the formula (Standard Deviation of measured concentration / Mean of measured concentration) X 100

**Table 3b.** Limit of Quantification (LOQ) of ORF1ab SARS-CoV-2 RT-dPCR assay quantified using Clarity Plus™. Three independent experimental runs were performed with a series of expected ORF1ab concentrations ranging from 0.125 to 1 copies/μl.

| Expected Concentration (Copies/μl) | *Measured Concentration Run 1* | *Measured Concentration Run 2* | *Measured Concentration Run 3* | *Mean of measured concentration (Copies/μl)* | *Relative Uncertainty<sup>a</sup> (%)* |
| --- | --- | --- | --- | --- | --- |
| 1 | 0.94 | 0.91 | 0.81 | 0.89 | 7.68 |
| 0.5 | 0.45 | 0.63 | 0.29 | 0.46 | 37.25 |
| 0.25 | 0.44 | 0.15 | 0.26 | 0.28 | 51.67 |
| 0.125 | 0.26 | 0.23 | 0.40 | 0.30 | 30.59 |
| No Template Control | 0.00 | 0.00 | 0.00 | 0.00 | - |

### 3.3 SARS-CoV-2 Assay limit of detection (LOD) on inactivated SARS-CoV-2 virus

The preliminary SARS-CoV-2 RT-dPCR assay LOD for detecting N1 and ORF1ab targets in AccuPlex™ SARS-CoV-2 RNA (Seracare) was first obtained by performing serial dilutions spanning 2, 4, 8, 16, and 32 times respectively in a series of triplicates. Prior to dilution, SARS-CoV-2 RNA was first extracted using EX3600 Automated Nucleic Acid Extraction System (Liferiver™) and quantified using Clarity Plus™, resulting in a measured RNA stock concentration of 1.8 copies/µl. From the results (Table 4a), it was revealed that Clarity Plus™ was able to provide a detection rate of 100% by successfully quantifying 3 out of 3 replicates for concentrations up to a dilution factor of 16 times for N1 (expected concentration of 0.1125 copies/ µl). With regards to ORF1ab, detection rate of 100% of dilution factor of 8 times (expected concentration of 0.225copies/ µl). Further analysis of N1 revealed that, at 16 times dilution, 2 out of the 3 replicates of N1 detected only 0.07 copies/µl which indicated that only one positive partition was present in the replicates. Therefore, the LOD was set at 8 times dilution for both targets (expected concentration of 0.225 copies/μl) due to the high probability of obtaining negative results if more than 3 replicates were performed at 16 times dilution.

**Table 4a.** Preliminary determination of detection limit (LOD) of the N1 SARS-CoV-2 RT-dPCR assay using AccuPlex™ SARS-CoV-2 RNA triplicates at expected concentration of 1.8, 0.9, 0.45, 0.225, 0.1125, 0.05625 copies/μl.

|  | **Dilution /**<br>**<u>Expected Concentration (Copies/μl)</u>** |  |  |  |  |  |
| --- | --- | --- | --- | --- | --- | --- |
|  | **0x**<br>1.8 | **2x**<br>0.9 | **4x**<br>0.45 | **8x**<br>0.225 | **16x**<br>0.1125 | **32x**<br>0.05625 |
| No template control |  |  |  |  |  |  |
| **<u>Measured Concentration (Copies/μl) in triplicates</u>** |  |  |  |  |  |  |
| 0.00 | 1.74 | 1.38 | 0.53 | 0.14 | 0.07 | 0.26 |
| 0.00 | 2.17 | 1.57 | 0.78 | 0.43 | 0.21 | 0.29 |
| 0.00 | 1.29 | 1.54 | 0.89 | 0.41 | 0.07 | **0.00** |
| **<u>Mean Concentration (Copies/μl)</u>** |  |  |  |  |  |  |
| 0.00 | 1.73 | 1.50 | 0.73 | 0.33 | 0.09 | 0.18 |

The results represent the individual measured concentration of three replicates within independent dilutions. Mean concentration represents the mean of three measured concentration replicates within independent dilutions.

**Table 4b.** Preliminary determination of detection limit (LOD) of the ORF1ab SARS-CoV-2 RT-dPCR assay using AccuPlex™ SARS-CoV-2 RNA triplicates at expected concentration of 1.8, 0.9, 0.45, 0.225, 0.1125, 0.05625 copies/μl.

|  | **Dilution /**<br>**<u>Expected Concentration (Copies/μl)</u>** |  |  |  |  |  |
| --- | --- | --- | --- | --- | --- | --- |
|  | **0x**<br>1.8 | **2x**<br>0.9 | **4x**<br>0.45 | **8x**<br>0.225 | **16x**<br>0.1125 | **32x**<br>0.05625 |
| No template control |  |  |  |  |  |  |
| **<u>Measured Concentration (Copies/μl) in triplicates</u>** |  |  |  |  |  |  |
| 0.00 | 1.60 | 0.72 | 0.53 | 0.34 | 0.00 | 0.13 |
| 0.00 | 1.66 | 1.09 | 0.39 | 0.28 | 0.21 | 0.00 |
| 0.00 | 1.78 | 1.00 | 0.41 | 0.14 | 0.22 | 0.00 |
| **<u>Mean Concentration (Copies/μl)</u>** |  |  |  |  |  |  |
| 0.00 | 1.68 | 0.97 | 0.44 | 0.25 | 0.14 | 0.04 |

The results represent the individual measured concentration of three replicates within independent dilutions. Mean concentration represents the mean of three measured concentration replicates within independent dilutions.

Sensitivity of the N1 SARS-CoV-2 assay was then verified on both the Clarity Plus™ dPCR (JN Medsys) and QuantStudio® 3 qPCR (Thermo Fisher Scientific) platforms. A series of 20 replicates were performed on both platforms with two RNA aliquots that were serially diluted 8 and 16 times to achieve expected concentrations of 0.225 copies/μl (LOD concentration), and 0.1125 copies/μl (2-fold dilution of LOD concentration) respectively. Data obtained from Table 5a indicated that at expected concentration of 0.225 copies/μl, the N1 SARS-CoV-2 assay on both platforms was able to quantify 20 out of 20 replicates providing a 100% detection rate. At a lower concentration of 0.1125 copies/μl, the RT-qPCR assay was only able to quantify 15 out of 20 replicates while the RT-dPCR assay was able to successfully detect 17 out of 20 replicates. This indicates that if the viral RNA concentration obtained from clinical samples are indeed as low as 0.1125 copies/μl, then the SARS-CoV-2 RT-dPCR assay will be able to successfully and accurately detect and quantify 85% of samples, while RT-qPCR only provides a 75% successful detection rate. Although ORF1ab was not compared across both platforms, results further support the LoD to be 0.225 cp/ μl. Based on the results shown above, the same SARS-CoV-2 assay demonstrated slightly higher sensitivity when performed on the Clarity Plus™ dPCR system as compared with the qPCR platform. Interestingly, this finding differs from some reports where dPCR using droplet-based systems demonstrated higher sensitivities than the comparator COVID-19 RT-qPCR assays [19-21]. These conclusions might need to be further assessed as the differences observed could plausibly be attributed to the variations in sample volume input and/or different assays used for the comparison studies.

**Table 5a.** Comparison of N1 SARS-CoV-2 assay on both dPCR (Clarity Plus™) and qPCR (QuantStudio® 3) platforms. A series of 20 replicates were performed on two RNA aliquots of 0.225 copies/μl (LOD) and 0.1125 copies/μl (2-fold dilution of LOD concentration).

| Expected Concentration<br>(Copies/μl) | Clarity Plus™ (N1) |  | QuantStudio® 3 (N1) |  |
| --- | --- | --- | --- | --- |
|  | 0.225 | 0.1125 | 0.225 | 0.1125 |
|  | <u>Measured Concentration (Copies/μl)</u> |  | <u>Measured Concentration (Cq)</u> |  |
| 1 | 0.13 | 0.07 | 33.05 | 35.66 |
| 2 | 0.34 | 0.07 | 33.40 | **Undetermined** |
| 3 | 0.19 | 0.13 | 33.79 | 35.38 |
| 4 | 0.68 | 0.21 | 35.00 | 33.52 |
| 5 | 0.35 | 0.07 | 31.80 | 35.85 |
| 6 | 0.36 | 0.14 | 32.53 | 34.06 |
| 7 | 0.41 | 0.20 | 32.63 | **Undetermined** |
| 8 | 0.28 | 0.42 | 32.63 | 32.55 |
| 9 | 0.22 | **0.00** | 32.64 | **Undetermined** |
| 10 | 0.35 | 0.14 | 32.20 | 32.64 |
| 11 | 0.34 | 0.32 | 32.79 | 33.59 |
| 12 | 0.36 | **0.00** | 32.63 | **Undetermined** |
| 13 | 0.27 | 0.07 | 32.02 | **Undetermined** |
| 14 | 0.08 | 0.13 | 32.57 | 34.17 |
| 15 | 0.34 | **0.00** | 32.30 | 35.62 |
| 16 | 0.51 | 0.19 | 32.89 | 32.59 |
| 17 | 0.34 | 0.27 | 32.31 | 35.92 |
| 18 | 0.33 | 0.07 | 32.89 | 31.44 |
| 19 | 0.20 | 0.13 | 33.44 | 34.61 |
| 20 | 0.28 | 0.28 | 32.54 | 35.91 |
| NTC | 0.00 | 0.00 | - | - |
| Detection Rate | 100% | 85% | 100% | 75% |

The results represent the measured concentration of 20 replicates at two independent concentrations for both Clarity Plus™ and QuantStudio® 3.

**Table 5b.** ORF1ab SARS-CoV-2 assay on dPCR (Clarity Plus™). A series of 20 replicates were performed on two RNA aliquots of 0.225 copies/μl (LOD) and 0.1125 copies/μl (2-fold dilution of LOD concentration).

| Expected Concentration (Copies/μl) | Clarity Plus™ (ORF1ab) |  |
| --- | --- | --- |
|  | 0.225 | 0.1125 |
| Measured Concentration (Copies/μl) |  |  |
| 1 | 0.07 | **0.00** |
| 2 | 0.21 | **0.00** |
| 3 | 0.32 | 0.13 |
| 4 | 0.45 | 0.21 |
| 5 | 0.14 | **0.00** |
| 6 | 0.07 | 0.07 |
| 7 | 0.20 | **0.00** |
| 8 | 0.14 | **0.00** |
| 9 | 0.15 | 0.14 |
| 10 | 0.35 | 0.21 |
| 11 | 0.27 | 0.06 |
| 12 | **0.00** | **0.00** |
| 13 | 0.40 | **0.00** |
| 14 | 0.31 | 0.13 |
| 15 | 0.14 | 0.07 |
| 16 | 0.08 | **0.00** |
| 17 | 0.14 | **0.00** |
| 18 | 0.33 | **0.00** |
| 19 | 0.26 | 0.07 |
| 20 | 0.14 | 0.21 |
| NTC | 0.00 | 0.00 |
| Detection Rate | 95% | 50% |

## 4. Conclusion

This study illustrates the capabilities and performance of a RT-dPCR assay in SARS-CoV-2 RNA detection and quantification. Coupled with the Clarity Plus™ dPCR system, this assay demonstrated excellent linearity across 6 orders of magnitude along its dynamic range for both SARS-CoV-2 targets, and successfully detected low-copy viral RNA samples with high inter- and intra-assay precision. In light of these advantages, RT-dPCR using Clarity Plus™ can potentially be employed in clinical settings as a molecular diagnostic tool for effective detection of SARS-CoV-2 in patients with low viral load and/or monitoring effectiveness of treatment through absolute quantitation of viral load. In addition, this technology can also be valuable for environmental/wastewater surveillance of COVID-19 where reliable viral load quantification tools are required. Detection of SARS-CoV-2 in environmental/wastewater samples is critical since it can provide early warning signs for disease transmission and outbreaks in the community.

## Data Availability

All data generated or analysed during this study are included in this published article (and its supplementary information files).

