## Supplementary Tables 1-4 for "Clarity Plus™ digital PCR: A novel platform for absolute quantification of SARS-CoV-2"

### Supplementary Data

**Supplementary Table 1:** SARS-CoV-2 RT-dPCR assay linearity study on Clarity Plus™ with three independent experimental runs in duplicates, using a series of expected N1 concentrations ranging from 0.02 to 30,000 copies/µl

| Expected Concentration (Copies/µl) | Number of Positive Partition | Total Partition Number | Ratio[a] | Measured Concentration 1[b] (Copies/µl) | Mean Measured Concentration 1 (Copies/µl) | Number of Positive Partition | Total Partition Number | Ratio[a] | Measured Concentration 2[b] (Copies/µl) | Mean Measured Concentration 2 (Copies/µl) | Number of Positive Partition | Total Partition Number | Ratio[a] | Measured Concentration 3[b] (Copies/µl) | Mean Measured Concentration 3 (Copies/µl) |
|---|---|---|---|---|---|---|---|---|---|---|---|---|---|---|---|
| 30000 | 41002 | 41064 | 0.99849 | 20954.04 | 22272.25 | 40527 | 40604 | 0.99810 | 20218.76 | 21418.38 | 44561 | 44627 | 0.99852 | 21020.77 | 20045.35 |
| | 44962 | 44992 | 0.99933 | 23590.46 | | 41011 | 41048 | 0.99910 | 22618.00 | | 46409 | 46535 | 0.99729 | 19069.93 | |
| 20000 | 42681 | 42811 | 0.99696 | 18700.05 | 18924.60 | 39414 | 39502 | 0.99777 | 19699.26 | 18887.03 | 46383 | 46515 | 0.99716 | 18918.48 | 18498.95 |
| | 42281 | 42393 | 0.99736 | 19149.16 | | 41084 | 41236 | 0.99631 | 18074.79 | | 43850 | 44012 | 0.99632 | 18079.42 | |
| 10000 | 41225 | 42829 | 0.96255 | 10595.85 | 10502.90 | 44826 | 46930 | 0.95517 | 10015.54 | 10048.33 | 45598 | 48068 | 0.94861 | 9575.48 | 9935.72 |
| | 43934 | 45749 | 0.96033 | 10409.95 | | 43744 | 45754 | 0.95607 | 10081.11 | | 43792 | 45669 | 0.95890 | 10295.95 | |
| 2000 | 21044 | 50136 | 0.41974 | 1755.73 | 1774.03 | 20235 | 46456 | 0.43557 | 1844.98 | 1827.02 | 21075 | 46281 | 0.45537 | 1960.16 | 1888.77 |
| | 20350 | 47738 | 0.42629 | 1792.33 | | 20274 | 47231 | 0.42925 | 1809.06 | | 20340 | 47223 | 0.43072 | 1817.38 | |
| 200 | 2791 | 45392 | 0.06149 | 204.70 | 205.88 | 2758 | 46164 | 0.05974 | 198.72 | 198.63 | 2772 | 47031 | 0.05894 | 195.96 | 2204.38 |
| | 2900 | 46642 | 0.06218 | 207.07 | | 2838 | 47545 | 0.05969 | 198.54 | | 2967 | 46475 | 0.06384 | 212.81 | |
| 20 | 296 | 43103 | 0.00687 | 22.23 | 22.39 | 298 | 43256 | 0.00689 | 22.30 | 21.55 | 302 | 42674 | 0.00708 | 22.91 | 22.16 |
| | 304 | 43612 | 0.00697 | 22.56 | | 250 | 38888 | 0.00642 | 20.80 | | 285 | 43102 | 0.00661 | 21.40 | |
| 2 | 22 | 42285 | 0.00052 | 1.68 | 1.79 | 30 | 45023 | 0.00067 | 2.15 | 2.48 | 32 | 49529 | 0.00065 | 2.08 | 1.75 |
| | 27 | 45717 | 0.00059 | 1.91 | | 39 | 44662 | 0.00087 | 2.82 | | 22 | 50111 | 0.00044 | 1.42 | |
| 1 | 13 | 43257 | 0.00030 | 0.97 | 0.97 | 16 | 45871 | 0.00035 | 1.13 | 1.32 | 14 | 42618 | 0.00033 | 1.06 | 1.06 |
| | 14 | 46659 | 0.00030 | 0.97 | | 19 | 40482 | 0.00047 | 1.51 | | 16 | 48865 | 0.00033 | 1.06 | |
| 0.2 | 3 | 47065 | 0.00006 | 0.21 | 0.31 | 1 | 44159 | 0.00002 | 0.07 | 0.11 | 3 | 45494 | 0.00007 | 0.21 | 0.21 |
| | 6 | 47428 | 0.00013 | 0.41 | | 2 | 42523 | 0.00005 | 0.15 | | 3 | 47429 | 0.00006 | 0.20 | |
| 0.02 | 3 | 45334 | 0.00007 | 0.21 | 0.17 | 3 | 41621 | 0.00007 | 0.23 | 0.15 | 1 | 45595 | 0.00002 | 0.07 | 0.10 |
| | 2 | 46210 | 0.00004 | 0.14 | | 1 | 44579 | 0.00002 | 0.07 | | 2 | 49740 | 0.00004 | 0.13 | |

The results represent individual positive signals, total partition number, ratio, and measured concentration of two replicates within independent experimental runs.

[a]Ratio was calculated by the formula (Number of Positive Signals / Total Partition Number)

[b]Measured Concentration (Copies/µl) is calculated by the formula: -ln(1- Ratio) / Partition Volume X 1000. Partition volume used is 0.31nL.

### Supplementary Data

**Supplementary Table 2:** SARS-CoV-2 RT-dPCR assay linearity study on Clarity Plus™ with three independent experimental runs in duplicates, using a series of expected ORF1ab concentrations ranging from 0.02 to 30,000 copies/µl

| Expected Concentration (Copies/µl) | Number of Positive Partition | Total Partition Number | Ratio[a] | Measured Concentration 1[b] (Copies/µl) | Mean Measured Concentration 1 (Copies/µl) | Number of Positive Partition | Total Partition Number | Ratio[a] | Measured Concentration 2[b] (Copies/µl) | Mean Measured Concentration 2 (Copies/µl) | Number of Positive Partition | Total Partition Number | Ratio[a] | Measured Concentration 3[b] (Copies/µl) | Mean Measured Concentration 3 (Copies/µl) |
|---|---|---|---|---|---|---|---|---|---|---|---|---|---|---|---|
| 30000 | 41034 | 41064 | 0.99927 | 23295.77 | 20274.11 | 40577 | 40604 | 0.99934 | 23599.31 | 22866.27 | 44570 | 44627 | 0.99872 | 21493.69 | 21324.76 |
| | 44778 | 44992 | 0.99524 | 17252.46 | | 41005 | 41048 | 0.99895 | 22133.22 | | 46469 | 46535 | 0.99858 | 21155.82 | |
| 20000 | 42566 | 42811 | 0.99428 | 16655.78 | 17067.08 | 39334 | 39502 | 0.99575 | 17613.36 | 17527.18 | 46195 | 46515 | 0.99312 | 16061.96 | 16326.37 |
| | 42205 | 42393 | 0.99557 | 17478.38 | | 41051 | 41236 | 0.99551 | 17441.00 | | 43755 | 44012 | 0.99416 | 16590.78 | |
| 10000 | 40203 | 42829 | 0.93869 | 9005.66 | 9020.12 | 43757 | 46930 | 0.93239 | 8690.26 | 8711.51 | 44570 | 48068 | 0.92723 | 8452.98 | 8579.86 |
| | 42969 | 45749 | 0.93923 | 9034.58 | | 42701 | 45754 | 0.93327 | 8732.76 | | 42597 | 45669 | 0.93273 | 8706.74 | |
| 2000 | 20021 | 50136 | 0.39933 | 1644.24 | 1662.72 | 19707 | 46456 | 0.42421 | 1780.67 | 1696.34 | 18825 | 46281 | 0.40675 | 1684.34 | 1684.34 |
| | 19390 | 47738 | 0.40618 | 1681.20 | | 18576 | 47231 | 0.39330 | 1612.01 | | 19208 | 47223 | 0.40675 | 1684.33 | |
| 200 | 2419 | 45392 | 0.05329 | 176.66 | 175.54 | 2457 | 46164 | 0.05322 | 176.43 | 172.23 | 2191 | 47031 | 0.04659 | 153.89 | 158.37 |
| | 2455 | 46642 | 0.05263 | 174.42 | | 2413 | 47545 | 0.05075 | 168.02 | | 2288 | 46475 | 0.04923 | 162.85 | |
| 20 | 262 | 43103 | 0.00608 | 19.67 | 20.30 | 250 | 43256 | 0.00578 | 18.70 | 17.66 | 239 | 42674 | 0.00560 | 18.12 | 17.09 |
| | 282 | 43612 | 0.00647 | 20.93 | | 200 | 38888 | 0.00514 | 16.63 | | 214 | 43102 | 0.00497 | 16.06 | |
| 2 | 17 | 42285 | 0.00040 | 1.30 | 1.43 | 29 | 45023 | 0.00064 | 2.08 | 2.05 | 17 | 49529 | 0.00034 | 1.11 | 1.30 |
| | 22 | 45717 | 0.00048 | 1.55 | | 28 | 44662 | 0.00063 | 2.02 | | 23 | 50111 | 0.00046 | 1.48 | |
| 1 | 16 | 43257 | 0.00037 | 1.19 | 0.97 | 6 | 45871 | 0.00013 | 0.42 | 0.81 | 13 | 42618 | 0.00031 | 0.98 | 1.09 |
| | 11 | 46659 | 0.00024 | 0.76 | | 15 | 40482 | 0.00037 | 1.20 | | 18 | 48865 | 0.00037 | 1.19 | |
| 0.2 | 3 | 47065 | 0.00006 | 0.21 | 0.28 | 1 | 44159 | 0.00002 | 0.07 | 0.19 | 2 | 45494 | 0.00004 | 0.14 | 0.58 |
| | 5 | 47428 | 0.00011 | 0.34 | | 4 | 42523 | 0.00009 | 0.30 | | 15 | 47429 | 0.00032 | 1.02 | |
| 0.02 | 1 | 45334 | 0.00002 | 0.07 | 0.18 | 2 | 41621 | 0.00005 | 0.16 | 0.19 | 6 | 45595 | 0.00013 | 0.42 | 0.21 |
| | 4 | 46210 | 0.00009 | 0.28 | | 3 | 44579 | 0.00007 | 0.22 | | 0 | 49740 | 0.00000 | 0.00 | |

The results represent individual positive signals, total partition number, ratio, and measured concentration of two replicates within independent experimental runs.

[a]Ratio was calculated by the formula (Number of Positive Signals / Total Partition Number)

[b]Measured Concentration (Copies/µl) is calculated by the formula: -ln(1- Ratio) / Partition Volume X 1000. Partition volume used is 0.31nL.

### Supplementary Data

**Supplementary Table 3:** Limit of Quantification (LOQ) in SARS-CoV-2 RT-dPCR assay quantified using Clarity Plus™. Three independent runs in duplicates were performed with a series of expected N1 concentrations ranging from 0.125 to 1 Copies/µl

| Expected Concentration (Copies/µl) | Number of Positive Partition | Total Partition Number | Ratio[a] | Measured Concentration 1[b] (Copies/µl) | Number of Positive Partition | Total Partition Number | Ratio[a] | Measured Concentration 2[b] (Copies/µl) | Number of Positive Partition | Total Partition Number | Ratio[a] | Measured Concentration 3[b] (Copies/µl) |
|---|---|---|---|---|---|---|---|---|---|---|---|---|
| 1 | 10 | 41267 | 0.00024 | 0.78 | 12 | 39938 | 0.00030 | 0.97 | 12 | 43297 | 0.00028 | 0.89 |
| | 16 | 44536 | 0.00036 | 1.16 | 14 | 41669 | 0.00034 | 1.08 | 15 | 47542 | 0.00032 | 1.02 |
| 0.5 | 6 | 41436 | 0.00014 | 0.47 | 7 | 45073 | 0.00016 | 0.50 | 7 | 43668 | 0.00016 | 0.52 |
| | 7 | 44060 | 0.00016 | 0.51 | 7 | 43235 | 0.00016 | 0.52 | 6 | 44058 | 0.00014 | 0.44 |
| 0.25 | 4 | 46397 | 0.00009 | 0.28 | 1 | 41806 | 0.00002 | 0.08 | 5 | 43130 | 0.00012 | 0.37 |
| | 3 | 42857 | 0.00007 | 0.23 | 2 | 43240 | 0.00005 | 0.15 | 1 | 44437 | 0.00002 | 0.07 |
| 0.125 | 3 | 42761 | 0.00007 | 0.23 | 3 | 42145 | 0.00007 | 0.23 | 3 | 44287 | 0.00007 | 0.22 |
| | 3 | 43267 | 0.00007 | 0.22 | 2 | 41699 | 0.00005 | 0.15 | 2 | 44112 | 0.00005 | 0.15 |

The results represent individual positive signals, total partition number, ratio, and measured concentration of two replicates within independent experimental runs.

[a]Ratio was calculated by the formula (Number of Positive Signals / Total Partition Number)

[b]Measured Concentration (Copies/µl) is calculated by the formula: -ln(1- Ratio) / Partition Volume X 1000. Partition volume used is 0.31nL

### Supplementary Data

**Supplementary Table 4:** Limit of Quantification (LOQ) in SARS-CoV-2 RT-dPCR assay quantified using Clarity Plus™. Three independent runs in duplicates were performed with a series of expected ORF1ab concentrations ranging from 0.125 to 1 Copies/µl

| Expected Concentration (Copies/µl) | Number of Positive Partition | Total Partition Number | Ratio[a] | Measured Concentration 1[b] (Copies/µl) | Number of Positive Partition | Total Partition Number | Ratio[a] | Measured Concentration 2[b] (Copies/µl) | Number of Positive Partition | Total Partition Number | Ratio[a] | Measured Concentration 3[b] (Copies/µl) |
|---|---|---|---|---|---|---|---|---|---|---|---|---|
| 1 | 15 | 41267 | 0.00036 | 1.17 | 8 | 39938 | 0.00020 | 0.65 | 17 | 43297 | 0.00039 | 1.27 |
| | 10 | 44536 | 0.00022 | 0.72 | 15 | 41669 | 0.00036 | 1.16 | 5 | 47542 | 0.00011 | 0.34 |
| 0.5 | 5 | 41436 | 0.00012 | 0.39 | 6 | 45073 | 0.00013 | 0.43 | 8 | 43668 | 0.00018 | 0.59 |
| | 7 | 44060 | 0.00016 | 0.51 | 11 | 43235 | 0.00025 | 0.82 | 0 | 44058 | 0.00000 | 0.00 |
| 0.25 | 4 | 46397 | 0.00009 | 0.28 | 2 | 41806 | 0.00005 | 0.15 | 2 | 43130 | 0.00005 | 0.15 |
| | 8 | 42857 | 0.00019 | 0.60 | 2 | 43240 | 0.00005 | 0.15 | 5 | 44437 | 0.00011 | 0.36 |
| 0.125 | 2 | 42761 | 0.00005 | 0.15 | 2 | 42145 | 0.00005 | 0.15 | 4 | 44287 | 0.00009 | 0.29 |
| | 5 | 43267 | 0.00012 | 0.37 | 4 | 41699 | 0.00010 | 0.31 | 7 | 44112 | 0.00016 | 0.51 |

The results represent individual positive signals, total partition number, ratio, and measured concentration of two replicates within independent experimental runs.

[a]Ratio was calculated by the formula (Number of Positive Signals / Total Partition Number)

[b]Measured Concentration (Copies/µl) is calculated by the formula: -ln(1- Ratio) / Partition Volume X 1000. Partition volume used is 0.31nL
